# Prognostic Value of Serum/Plasma Neurofilament Light Chain for COVID-19 Associated Mortality

**DOI:** 10.1101/2022.01.13.22269244

**Authors:** Ruturaj R. Masvekar, Peter Kosa, Kimberly Jin, Kerry Dobbs, Michael A. Stack, Riccardo Castagnoli, Virginia Quaresima, Helen C. Su, Luisa Imberti, Luigi D. Notarangelo, Bibiana Bielekova

## Abstract

Given the continued spread of severe acute respiratory syndrome coronavirus 2 (SARS-CoV-2), early predictors of coronavirus disease 19 (COVID-19) mortality might improve patients’ outcomes. Increased levels of circulating neurofilament light chain (NfL), a biomarker of neuro-axonal injury, have been observed in patients with severe COVID-19. We investigated whether NfL provides non-redundant clinical value to previously identified predictors of COVID-19 mortality.

We measured serum or plasma NfL concentrations in a blinded fashion in 3 cohorts totaling 338 COVID-19 patients. In cohort 1, we found significantly elevated NfL levels only in critically ill COVID-19 patients compared to healthy controls. Longitudinal cohort 2 data showed that NfL is elevated late in the course of the disease, following two other prognostic markers of COVID-19: decrease in absolute lymphocyte count (ALC) and increase in lactate dehydrogenase (LDH). Significant correlations between LDH and ALC abnormalities and subsequent rise of NfL implicate multi-organ failure as a likely cause of neuronal injury at the later stages of COVID-19. Addition of NfL to age and gender in cohort 1 significantly improved the accuracy of mortality prediction and these improvements were validated in cohorts 2 and 3.

In conclusion, although substantial increase in serum/plasma NfL reproducibly enhances COVID-19 mortality prediction, NfL has clinically meaningful prognostic value only close to death, which may be too late to alter medical management. When combined with other prognostic biomarkers, rising longitudinal NfL measurements triggered by LDH and ALC abnormalities would identify patients at risk of COVID-19 associated mortality who might still benefit from escalated care.

## INTRODUCTION

Since early 2020, the COVID-19 pandemic has exhausted medical systems worldwide. Even after the development of safe and effective vaccines, SARS-CoV-2 continues to spread (COVID Live Update - Worldometer). A reliable early predictor of COVID-19 associated mortality would help prioritize use of medical resources and maximize patient survival.

Neurofilaments are essential cytoskeleton proteins of the central and peripheral axons exclusive to the nervous system. Of three neurofilament subunits, neurofilament light chain (NfL) has the lowest molecular weight and easily diffuses from parenchyma to CSF and blood (Fuchs and Cleveland, 1998; Scherling et al., 2014; Alirezaei et al., 2020). Recent developments of ultrasensitive assays, such as Single Molecule Array (SIMOA), allow reproducible measurement of low NfL concentrations in serum or plasma (Rissin et al., 2010; Kan et al., 2012). Consequently, blood NfL became a key noninvasive biomarker of acute neuronal injury in diverse neuropathological conditions (Barro et al., 2020).

Although previous studies have demonstrated association between COVID-19 morbidity and CNS damage (Aamodt et al., 2020; Ameres et al., 2020; Kanberg et al., 2020, 2021; Prudencio et al., 2021), several questions still remain unanswered: 1) Does a single measurement of NfL provide meaningful prognostic information at individual patient level?; 2) Is there a relationship between NfL and previously described COVID-19-associated mortality biomarkers (Yan et al., 2020) of prognostic value, such as ALC, C-reactive protein [CRP] and LDH?; and 3) Does NfL improve COVID-19 mortality prediction by demographic markers such as age and gender?

## MATERIALS and METHODS

### Research subjects and cohorts

Serum or plasma samples from COVID-19 patients admitted at ASST Spedali Civili (Brescia, Italy) were obtained through Laboratory of Clinical Immunology and Microbiology (LCIM), National Institute of Allergy and Infectious Diseases (NIAID), under Institutional Review Board (IRB)-approved protocols (Comitato Etico Provinciale: NP 4000 - Studio CORONAlab and NP 4408 - Studio CORONAlab and ClinicalTrials.gov: NCT04582903). SARS-CoV-2 infection was confirmed using nasopharyngeal swab – polymerase chain reaction (PCR) test. COVID-19 disease severity was determined as per Diagnosis and Treatment Protocol for Novel Coronavirus Pneumonia guidelines, released by the National Health Commission & State Administration of Traditional Chinese Medicine (Wei PF, 2020). Serum and plasma samples from healthy controls (HC) and multiple sclerosis (MS) subjects were collected at Neuroimmunological Diseases Section (NDS), NIAID after informed consent under IRB-approved protocol (ClinicalTrials.gov: NCT00794352). The NfL levels measured in HC and MS subgroups were previously reported (Masvekar R et al., 2021) and are used in the current study only as a positive control of neuronal injury; the measurements of other COVID-19 prognostic biomarkers in these control samples were not reported previously.

378 serum or plasma samples were collected from 338 COVID-19 patients grouped into 3 independent cohorts (Figure 1, Table 1, and Supplementary Data File 1). In cohort 1, 30 cross-sectional samples were collected from COVID-19 patients with 3 levels of disease severity. In cohort 2, 60 longitudinal samples were collected from 20 critically ill COVID-19 patients (T1, T2, and T3: collected averagely at 5 to 10 day intervals, within 30 days of hospitalization). Cohort 3 consisted of 288 cross-sectional samples collected from critically ill COVID-19 patients where a large proportion of the subjects eventually died (39.2%).

**Table 1:**
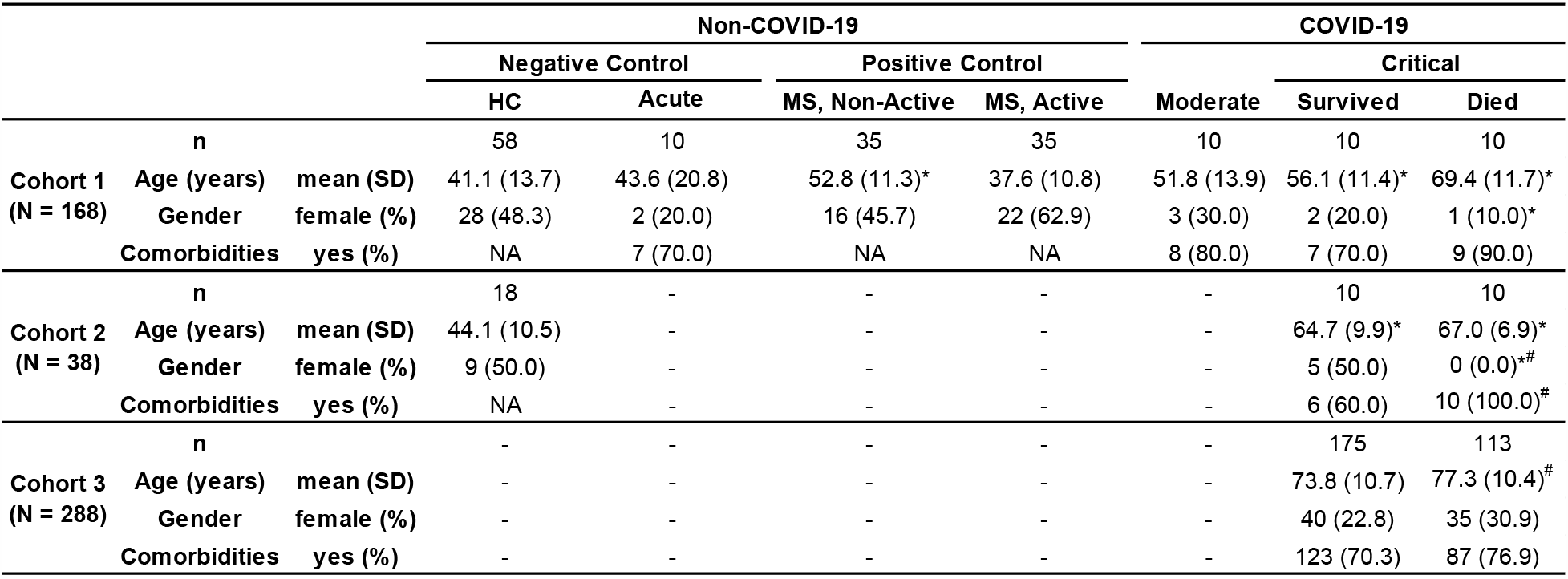
Demographic details of the 3 cohorts. Age (ANOVA or unpaired t-test), gender, and comorbidities distribution) were compared across disease diagnosis and severity subgroups using Chi-square test. *p < 0.05 vs HC and ^#^p < 0.05 vs COVID-19, Critical - Survived.

**Figure 1:**
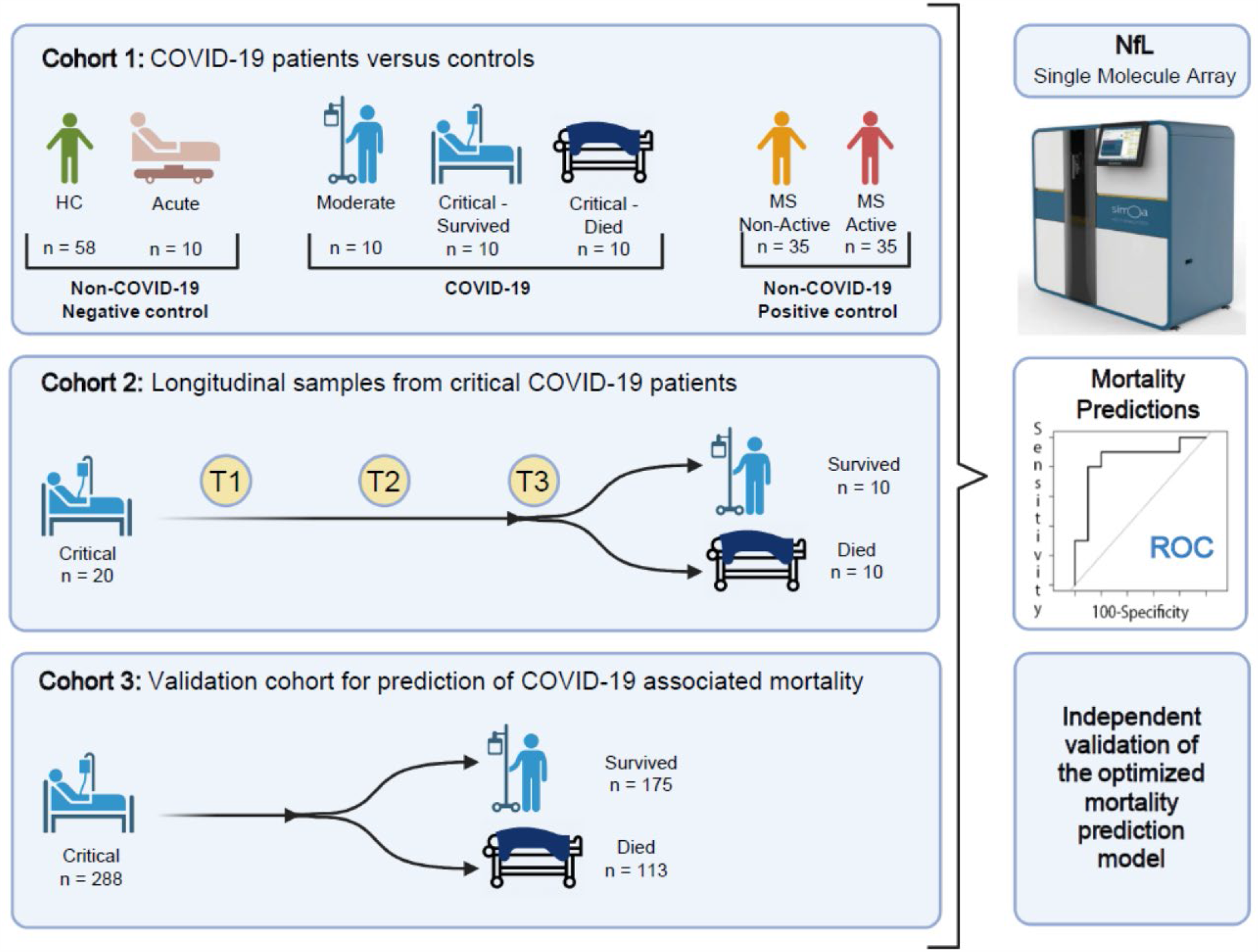
Patient selection, objectives, and experiment outlines of 3 independent cohorts. Cohort 1 aims to analyze NfL cross-sectionally across disease diagnosis and severity categories. In cohort 2, objective was to analyze NfL levels in critically ill COVID-19 patients, longitudinally at 3 different time-points (T1, T2, and T3: collected averagely at 5 to 10 days interval, within 30 days of hospitalization). Observed additional prognostic value of NfL with traditional demographic factors (age and gender) from cohorts 1 and 2, was independently validated in cohort 3.

### NfL single molecular array (Simoa™) assay

NfL concentrations in serum or plasma samples were measured using Simoa™ assay (Catalog # 103186; Quanterix, Billerica, MA, USA). Samples were diluted 1:4 and randomly distributed on 96-well plates. Quality control (QC) samples provided with the kit had concentrations within the pre-defined range and the coefficient of variance (CV) across the plates was < 10%. All samples were analyzed blindly under alpha-numeric codes. The diagnostic codes were broken only after QC verified NfL concentrations were reported to the database manager.

### Adjustment for effect of healthy aging

As serum/plasma NfL levels increase with physiological aging (Disanto et al., 2017), the measured NfL concentrations were adjusted for effect of healthy aging as described previously (Masvekar R et al., 2021). Following age vs serum- or plasma-NfL equations from HC cohorts were used: ln(serum NfL) = 0.0177*Age + 0.9696 and ln(plasma NfL) = 0.0158*Age + 1.247. The age-adjusted NfL concentrations represent residuals from the above-stated linear regression models.

### Statistical analyses

NfL levels were compared across disease diagnosis and severity subgroups using either Kruskal-Wallis ANOVA or Welch’s t-test. Correlations between NfL and systemic markers of COVID-19 morbidity were evaluated using Spearman analysis and linear regression model.

Prediction models of COVID-19 associated mortality were developed in R Studio Version 1.1.463 (R version 4.0.2) using logistic regression (*glm* function of the “stat” package) (R: The R Project for Statistical Computing). Optimal cutoff for the predictive models was calculated using *optimalCutoff* function of the “InformationValue” package (https://cran.r-project.org/web/packages/InformationValue/index.html). The receiver operating characteristic curve (ROC) was calculated using *roc* function of the “pROC” package (Robin et al., 2011).

## RESULTS

### NfL levels increase with COVID-19 severity and mortality

Although increased blood NfL levels have been reported in patients with severe COVID-19 (Aamodt et al., 2020; Ameres et al., 2020; Kanberg et al., 2020, 2021; Prudencio et al., 2021), previous studies had insufficient numbers of subjects who died from the disease to assess whether NfL can predict COVID-19 mortality.

To fill this knowledge gap, we measured NfL levels in 30 COVID-19 patients with 3 levels of severity: 1) moderate severity (n = 10); 2) critical condition but survived (n = 10); and 3) critical condition but died (n = 10). Positive and negative control subgroups consisted of 1) patients with acute COVID-19-like symptoms admitted in critical health conditions who tested negative for SARS-CoV-2 infection (n = 10); 2) HC (n = 58); 3) MS patients with acute focal CNS inflammation measured as contrast-enhancing lesions on brain MRI (active MS, n = 35); and 4) MS patients without evidence of acute focal CNS inflammation (non-active, n = 35).

After diagnostic codes were unblinded, we found elevated levels of NfL in COVID-19 patients compared to HC (Figure 2A). NfL levels in COVID-19 patients increased with disease severity, but only cohorts of critically ill COVID-19 and MS patients reached statistical significance compared to HC.

**Figure 2:**
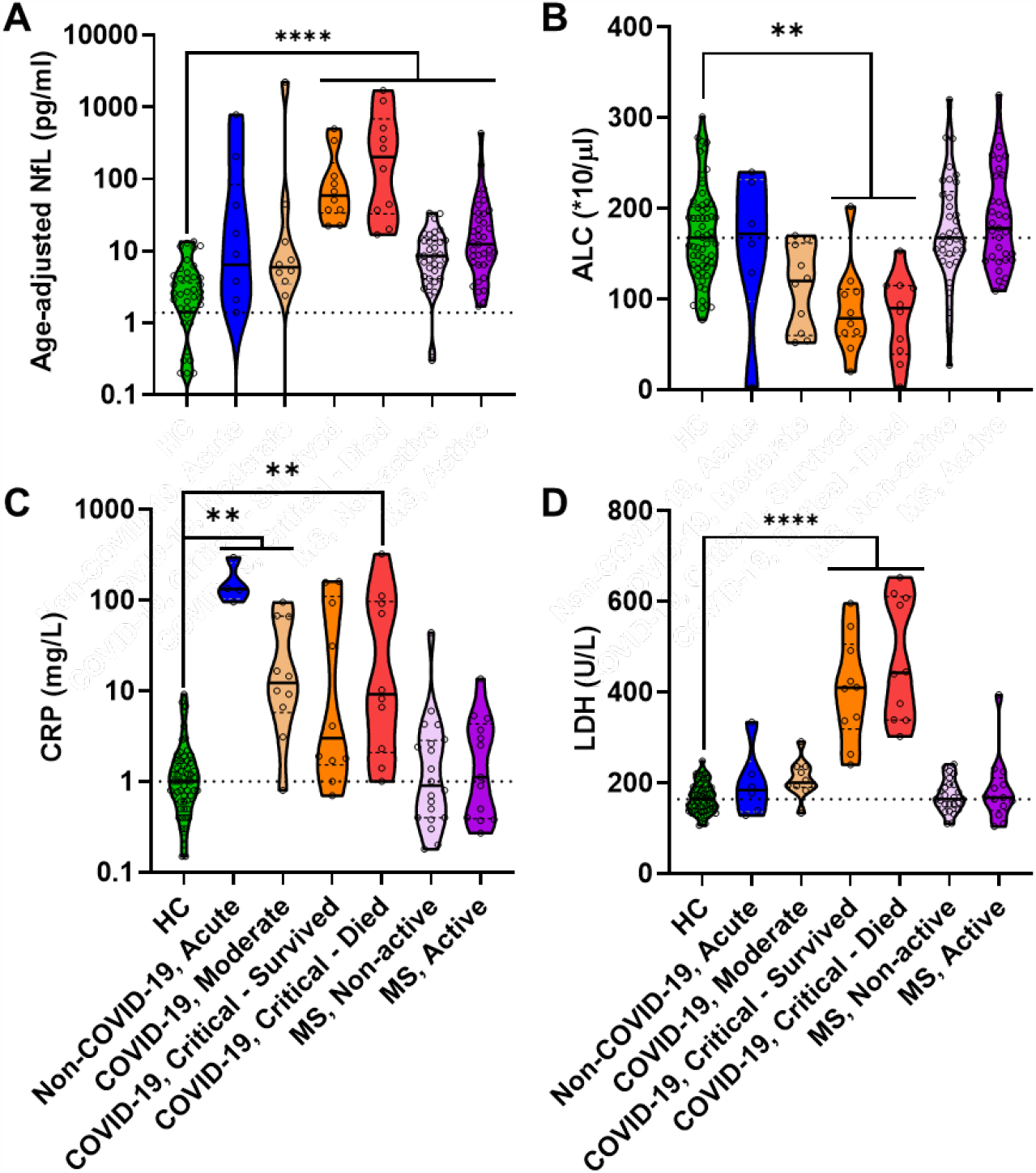
In cohort 1, **(A)** NfL, **(B)** ALC, **(C)** CRP, and **(D)** LDH were compared across HC vs. COVID-19 disease severity and multiple sclerosis disease activity subgroups using Kruskal-Wallis ANOVA; **p < 0.005 and ****p < 0.0001. The dotted line on each plot indicates the median of HC.

Next, we compared cohort differences in other blood biomarkers of COVID-19 morbidity: ALC, CRP, and LDH (Figures 2B, 2C and 2D). Like NfL, decreased ALC and increased LDH correlated with COVID-19 severity; statistically significant differences in ALC and LDH were observed only in critically ill COVID-19 patients compared to HC. Interestingly, although non-COVID-19 acute respiratory illness control had levels of COVID-19 prognostic biomarkers (i.e., NfL, ALC, and LDH) comparable to HC, they had the highest CRP levels.

We conclude that NfL, LDH, and ALC abnormalities increase with COVID-19 severity, are associated with COVID-19 mortality, and can differentiate COVID-19 from other acute respiratory conditions that lead to ICU admission.

### In COVID-19 patients NfL rises close to death, trailing transient abnormalities in ALC and LDH by 5 to 20 days

The earlier a biomarker can identify patients at risk for COVID-19 mortality, the greater its clinical value. Because none of the previous studies addressed the dynamics of NfL rise in COVID-19 and compared it to the dynamics of other prognostic biomarkers, we addressed this knowledge gap in the longitudinal cohort 2.

We measured NfL in 60 samples collected from 20 critically ill COVID-19 patients within 30 days of hospitalization, at three timepoints (T1, T2 and T3) taken at approximately 5 to 10 day intervals. We observed statistically significant, progressive increases (T1 vs. T2 and T3) in NfL levels only in patients who later died (Figure 3A).

**Figure 3:**
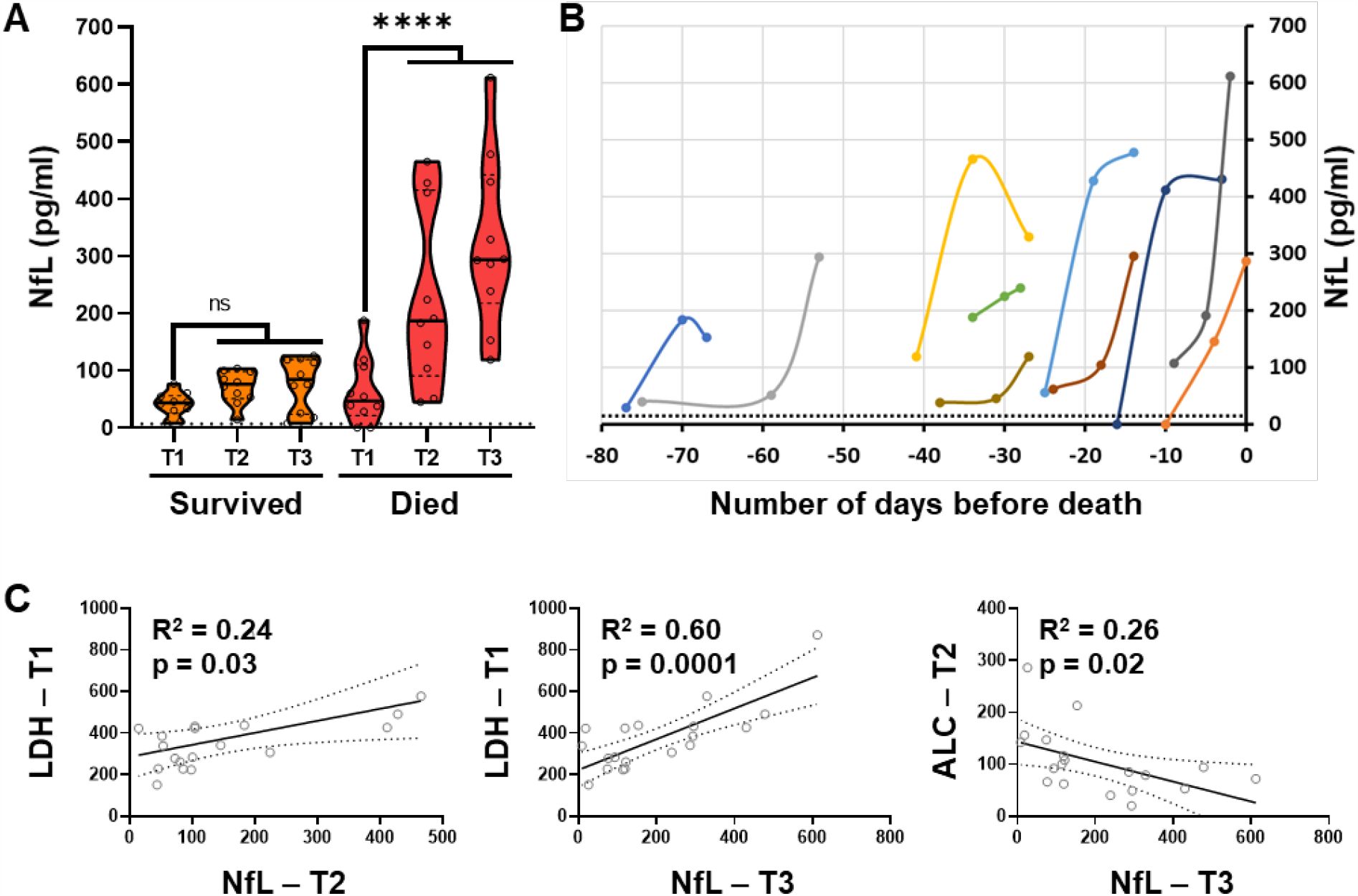
In cohort 2, **(A)** NfL levels at 3 different time points (T1, T2, and T3: collected on average at 5 to 10 day intervals, within 30 days of hospitalization) in critically ill COVID-19 patients were compared (survived versus died) using Kruskal-Wallis ANOVA; ****p < 0.0001. The dotted line indicates the median of the HC. **(B)** Longitudinal NfL levels in critical COVID-19 patients who died, plotted with respect to number of days before death. Each line represents data from an individual patient. The dotted line represents upper limit in HC (i.e., mean + 3*SD = 20 pg/ml). **(C)** Correlations between systemic biomarkers’ measurements at earlier time points (T1 and 2) and NfL measurements at later time points (T2 and T3) were assessed using linear regression analysis. R^2^ and p-value are represented on respective correlation plots. The dotted line indicates 95% confidence interval.

When plotting measurements against day of hospitalization, the greatest rise in NfL occurred close to death (Figure 3B and Supplementary Figure 1). Consistent with prior reports that NfL levels remain elevated for weeks (up to 3 months) following acute CNS injury (Thelin et al., 2017), increased NfL in COVID-19 patients did not return to normal within the observation period. In contrast, ALC, LDH, and CRP (Supplementary Figure 1) demonstrated large day-to-day fluctuations and were also frequently elevated in surviving patients (Supplementary Figure 2).

To assess if transient abnormalities in LDH, CRP, and ALC levels precede increases in NfL, we investigated correlations between these systemic markers measured at initial time-points (T1 and T2), with NfL measured later (i.e., T1 vs T2, T1 vs T3 and T2 vs T3). Only 3 of these comparisons reached statistical significance (Figure 3C), with the strongest relationship observed between LDH measured at first time point (T1) and NfL measured at last time point (T3), which explains almost 60% of variance (R^2^ = 0.598, p = 0.0001). Consistent with the lack of association of CRP measurements with COVID-19 severity, CRP elevations did not predict subsequent rise in NfL.

We conclude that critically ill COVID-19 patients experience earlier abnormalities in ALC and LDH measurements, which are strongly associated with later elevation in NfL levels. While these critically high NfL levels predict COVID-19 mortality, they peak shortly before death, which may be too late to alter medical management.

### NfL measured close to death enhances mortality prediction of age and gender-based classifier

As all the above-described observations supported clinical value of NfL to predict COVID-19 mortality, we sought to quantify this predictive value on an individual patient level and compare it to demographic prognostic markers such as age, gender, and comorbidities.

In the cohort 1, used as a training cohort, we predicted COVID-19 mortality using measured NfL as a continuous variable (Figure 4A, left panel). Single, cross-sectional NfL measurements could not reliably predict death, reaching an area under receive operator characteristic curve (AUROC) of only 0.61 with 95% confidence interval ([CI]: 0.33-0.89) crossing the value of random guessing (i.e., AUROC 0.5). The optimal cut-off from NfL to predict mortality from cohort 1 ROC curve was 124 pg/ml.

**Figure 4:**
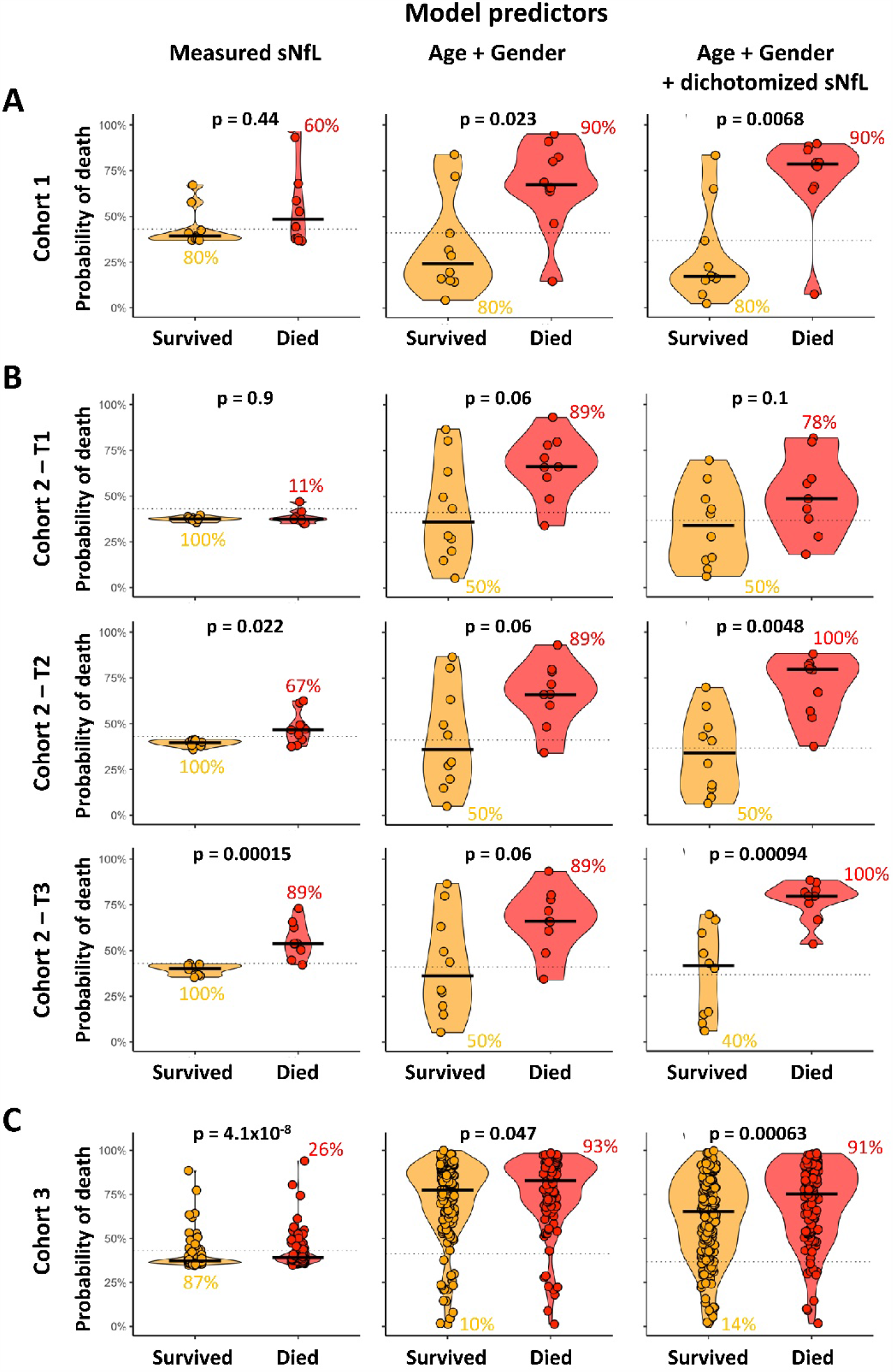
Comparisons of 3 predictive models of COVID-19 associated mortality: continuous NfL measurement, Age plus Gender, and Age plus Gender plus dichotomized NfL in 3 independent cohorts; **(A)** cohort 1, **(B)** cohort 2, and **(C)** cohort 3. The dotted line on each plot represents the optimal cut-off for respective model predictor. The numerical values beside respective subgroups (Survived or Died) on each plot represents the percentage of correctly classified patients.

As shown in Table 1, cohorts 1 and 2 were not matched for demographic predictors of COVID-19 mortality: in both cohorts, patients who survived were generally younger, with higher proportion of females and lower proportion of subjects with comorbidities. Therefore, it should not be surprising that NfL measurements alone, ignoring these important demographic variables, had low predictive power. Instead, we built a prognostic classifier that integrated NfL (dichotomized based on optimal cut-off 124 pg/ml) with age and gender, and compared it to the model(s) without NfL. We also tested a more complex classifier consisting of dichotomized NfL, age, gender, and comorbidities, but observed weaker independent validation of this model compared to a model without comorbidities (Supplementary Figure 3). For the sake of space and clarity we will present data only on the strongest model.

Adding dichotomized NfL enhanced predictive value of age and gender in cohort 1 from AUROC 0.8 to 0.85 and p-value from 0.023 to 0.0068 (Figure 4, cohort 1 panel).

Next, we sought to assess performance of the leading mortality predictor in Cohort 2, which did not contribute to model generation (Figure 4B). Addition of dichotomized NfL to the age and gender at first longitudinal time-point (T1) did not improve predictive value of the model, consistent with observation that at early timepoint the NfL values were indistinguishable between patients who survived and those who died. In contrast, NfL significantly improved the predictive power of the combined classifier at later time-points (T2 and T3; T2: AUROC from 0.76 (CI: 0.53-0.99) to 0.89 (CI: 0.74-1.00) and p-value from 0.06 to 0.0048; T3: AUROC from 0.76 (0.53-0.99) to 0.96 (0.87-1.00) and p-value from 0.06 to 0.00094).

We conclude that NfL measurement provides additive COVID-19 mortality predictive value to the traditional demographic prognostic factors, provided that NfL is measured in critically ill patients later in disease.

Finally, we were able to assess the non-redundant prognostic value of NfL in a unique large cohort of patients with high COVID-19 mortality risk (i.e., elderly patients with high proportion of males with comorbidities; Figure 4C). As expected, out of these 288 critically ill COVID-19 patients, a large proportion (n=113; 39.2%) eventually died.

Although surviving and dying cohorts were matched for age, gender, and comorbidities as univariate predictors (Table 1), the combined age plus gender model correctly predicted a marginally higher mortality in the cohort of subjects who eventually died (10% vs 93%; p=0.047). NfL levels differentiated survivors from non-survivors with much stronger statistical significance (p = 4.1e-08). Adding dichotomized NfL to demographic data improved the accuracy of mortality prediction compared to demographic data alone. Specifically, the AUROC increased from 0.57 (CI: 0.50-0.64) to 0.62 (CI: 0.55-0.69) and p-value improved from 0.047 to 0.00063. Nevertheless, the sensitivity (71.4%) and specificity (40.7%) of this predictor remained weak in this unique cohort.

## DISCUSSION

This study validates reports linking high serum/plasma NfL levels to COVID-19 severity (Aamodt et al., 2020; Ameres et al., 2020; Kanberg et al., 2020, 2021; Prudencio et al., 2021; Sutter et al., 2021). Our longitudinal measurements demonstrated that rise in NfL generally occurs during hospitalizations of critically ill patients and trails other transient laboratory abnormalities such as decreased ALC and increased LDH by 5 to 20 days. The degree of LDH increase is a strong determinant of subsequent magnitude of NfL rise, suggesting that COVID-19-associated CNS injury is secondary to damage of other critical organs, such as liver, kidneys, and lungs. This conclusion aligns with pathology studies ruling out strong primary infiltration of CNS tissue by the SARS-CoV-2 or by immune system; those studies instead attribute COVID-19 associated CNS damage to processes such as hypoxia or intravascular coagulation (Serrano et al., 2021).

Compared to previous studies of NfL in COVID-19, we studied a cohort of patients in which a high proportion eventually died (133/338 = 39.3%). This allowed us to unequivocally link high serum/plasma NfL levels also with COVID-19 mortality, something that remained ambiguous in the previous studies.

We constructed a model that combined demographic predictors of COVID-19 mortality with NfL measurement and validated its greater predictive accuracy. Nevertheless, the accuracy of this classifier varied between the cohorts, depending on the timing of NfL measurement (i.e., later measurements enhanced predictive power) and underlying premorbid risk. Indeed, comparing model performance among our 3 cohorts, it appeared that NfL has greater predictive value in younger (cohorts 1 and 2) versus older (cohort 3) subjects. This is perhaps not surprising as younger patients with fewer comorbidities have higher likelihood of withstanding multi-organ failure and therefore CNS injury may become key determinant of their survival. In contrast, elderly subjects with high premorbid risk rapidly succumb to multi-organ failure before CNS injury manifests clinically or by high NfL concentrations.

Integrating all our observations, we recommend that NfL should be measured longitudinally and integrated with existing prognostic markers to optimize care. For example, a screening NfL measurement at the beginning of hospitalization, expected to be normal in most patients, might identify a few subjects with either neurological comorbidity or with advanced stage of COVID-19 who require care focused on preventing further CNS injury. After an initial negative NfL test, critically ill COVID-19 patients might be best monitored by standard laboratory tests such as LDH and ALC. Identified spikes should prompt more aggressive management that includes longitudinal NfL monitoring approximately every 5 days. Any increase in NfL should be considered a poor prognostic indicator necessitating escalation therapies. These may include neuro-protective strategies that lower CNS metabolism, such as systemic cooling or barbiturates. Stabilization of NfL levels indicates that escalation therapy worked, while further increases signify continuous neuro-axonal injury that must be stopped to limit mortality.

While the COVID-19 pandemic demonstrated prognostic value of NfL in critically ill patients with SARS-CoV-2 infection, non-invasive, ultrasensitive measurement of NfL could be used to monitor neuronal injury in all comatose, or heavily sedated critically ill patients regardless of SARS-CoV-2 infection status. Ultra-sensitive assays will hopefully become broadly adopted by clinical laboratories and might include in the future other CNS-derived analytes for enhanced accuracy of non-invasive monitoring of CNS tissue.

## Data Availability

All data produced in the present study are available upon reasonable request to the authors.

## ACKNOWLEDGEMENTS

This study was supported by the Intramural Research Program of NIAID, NIH and by Regione Lombardia, Italy (project “Risposta immune in pazienti con COVID-19 e co-morbidità”). We thank additional personnel in the ASST Spedali Civili, Brescia, Italy and LCIM, NIH, NIAID for providing us COVID-19 patients’ serum/plasma samples.

## SUPPLEMENTARY FIGURES

**Supplementary Figure 1:**
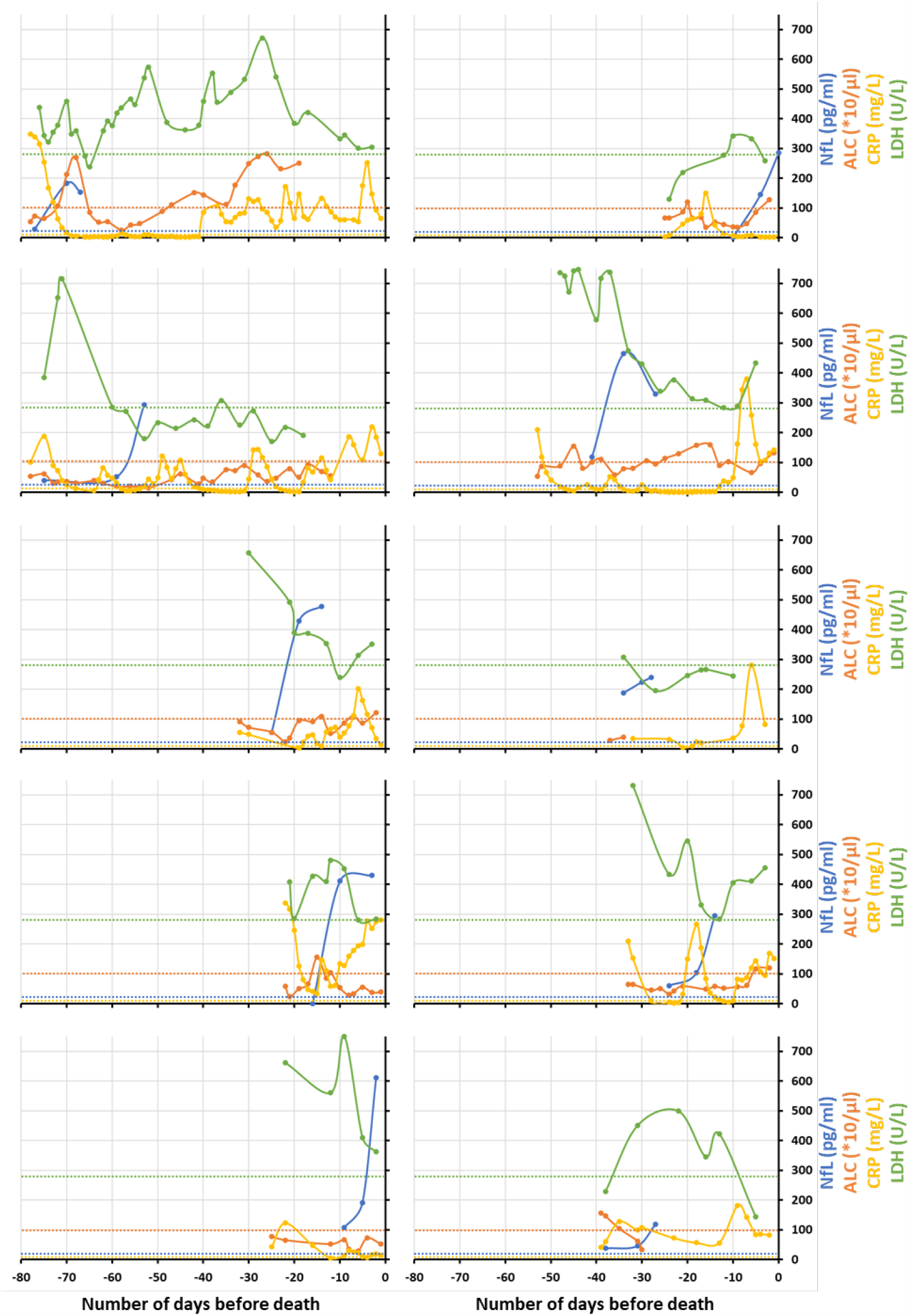
In cohort 2, longitudinal NfL (blue), ALC (orange), CRP (yellow) and LDH (green) levels in critically ill COVID-19 patients those who died, plotted with respect to number of days before death. Each plot represents an individual patient data. The respective color dotted lines represent upper (for NfL, CRP and LDH) or lower (for ALC) limit for HC for respective biomarker (NfL: 20 pg/ml, ALC: 100 *10/μl, CRP: 5 mg/L and LDH: 280 U/L).

**Supplementary Figure 2:**
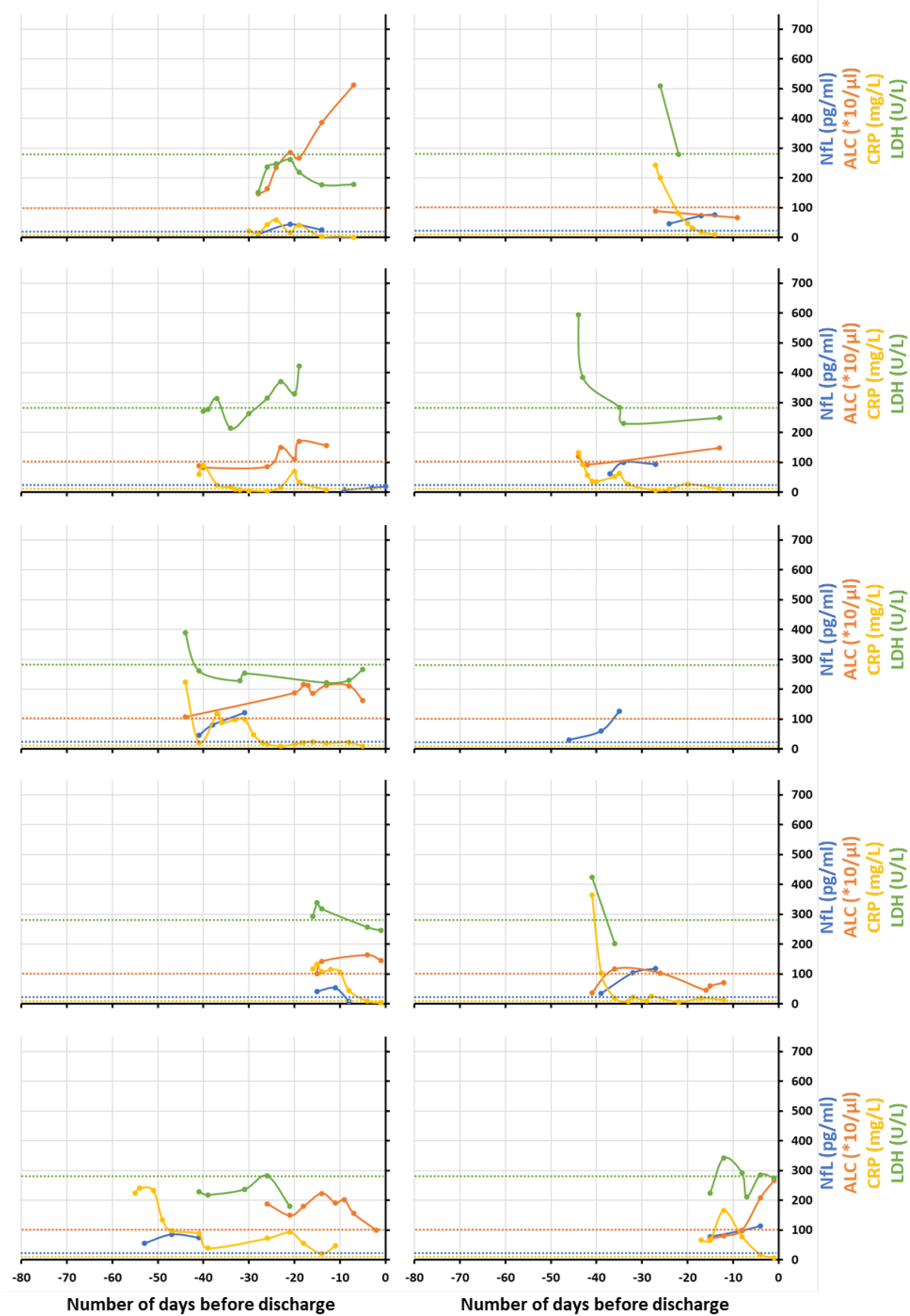
In cohort 2, longitudinal NfL (blue), ALC (orange), CRP (yellow) and LDH (green) levels in critically ill COVID-19 patients those who survived, plotted with respect to number of days before discharge. Each plot represents an individual patient data. The respective color dotted lines represent upper or lower limit for HC for respective biomarker.

**Supplementary Figure 3:**
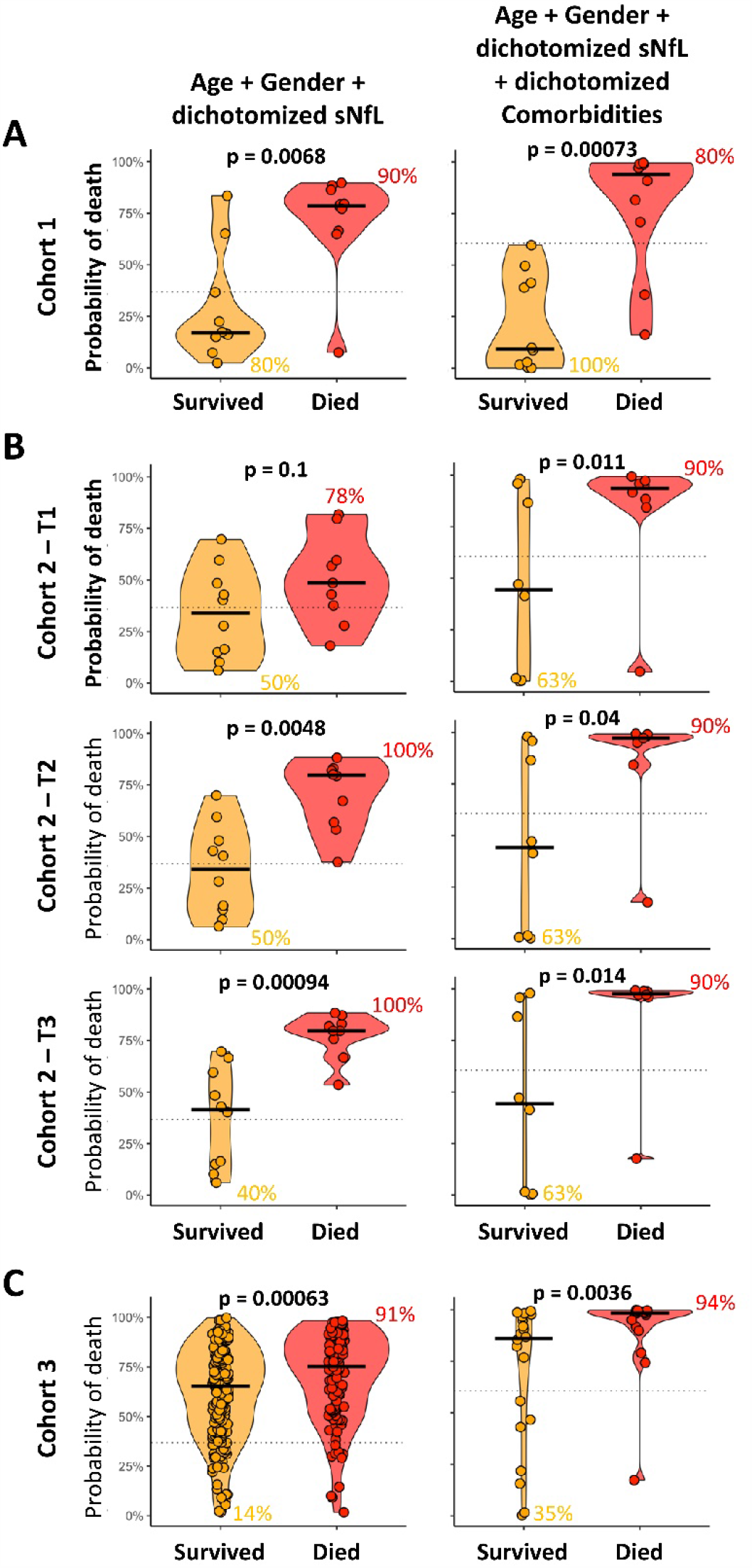
Comparisons of 2 predictive models of COVID-19 associated mortality: Age plus Gender plus dichotomized NfL and Age plus Gender plus dichotomized NfL plus dichotomized comorbidities in 3 independent cohorts; **(A)** cohort 1, **(B)** cohort 2, and **(C)** cohort 3. The dotted line on each plot represents the optimal cut-off for respective model predictor. The numerical values beside respective subgroups (Survived or Died) on each plot represents the percentage of correctly classified patients.

## SUPPLEMENTARY DATA FILE

**Supplementary data file 1:** Cohort, demographics, disease and severity diagnosis, timeline of important events during disease, NfL – raw and HC age-adjusted measurements, comorbidities and lab test measurements for systemic markers (ALC, CRP and LDH) data for all subjects (HC = 76, Non-COVID-19 Acute = 10, MS Non-active = 35, MS Active = 35, COVID-19: moderate = 10, critical - survived = 195, and critical - deceased = 133). Patients were recoded and personally identifiable information were excluded.

